# Perceptions and responses to COVID-19 through wastewater surveillance information and online search behavior: A randomized controlled trial

**DOI:** 10.1101/2023.10.20.23297297

**Authors:** Michio Murakami, Shuhei Nomura, Hiroki Ando, Masaaki Kitajima

## Abstract

Wastewater surveillance is anticipated to be a representative and timely method to assess infectious disease status; however, its influence on public perception and behavior remains unclear. Therefore, in this study, we used a randomized controlled trial to analyze the influence of wastewater surveillance-based information on understanding of, interest in, relief regarding, preventive behavioral intention against, and subsequent online search behavior related to coronavirus disease 2019 (COVID-19). Valid responses were obtained from 1,000 individuals in both control and intervention groups from Yahoo crowdsourcing users aged ≥18 years in Japan. This survey was conducted from August 4 to August 7, 2023, just before the common Japanese tradition of returning to hometowns. The questionnaire not only collected personal attributes but also gauged responses to COVID-19 information. This information highlighted the early detection capabilities and representativeness of wastewater surveillance compared with sentinel surveillance at medical institutions. At one-week post-survey, we obtained the survey participants’ online search history for key words such as “bullet train,” “highway,” “airplane,” and “wastewater.” The findings showed no significant differences between the two groups in terms of COVID-19 interest or preventive behavior before information provision, verifying the effectiveness of participant randomization. Wastewater surveillance-based information did not notably elevate understanding or specific intentions regarding COVID-19, such as wearing masks and receiving vaccination. However, it significantly increased interest in, relief concerning the infection status, and general preventive behavioral intentions. Heightened interest and general preventive intentions did not depend on prior interest or behavior. However, those who previously engaged in preventive behavior or who were less interested in COVID-19 exhibited more relief after exposure to wastewater surveillance-based information. Furthermore, this information could slightly influence online searches related to return travel modes, such as highways. In conclusion, information from wastewater surveillance effectively shapes individual perceptions of and responses to infections.

## 1. Introduction

During an infectious disease outbreak, it is vital to monitor infection status over time, including tracking the daily number of infected individuals. Monitoring helps ensure adequate medical resources, such as hospital capacity (Emanuel et al., 2020). Furthermore, sharing information about the infection status can prompt the public to adopt preventive behavior (Morii et al., 2023). Since the onset of the coronavirus disease 2019 (COVID-19) pandemic, data on daily infections have been widely disseminated in many countries (Mathieu et al., 2020). However, as the severity of the COVID-19 pandemic has decreased, obtaining a comprehensive picture of the infection status has become challenging. Comprehensive or random sampling surveys can be costly, and some symptomatic individuals avoid seeking medical attention (Dean, 2022).

Measuring pathogenic microorganisms in the influent of wastewater treatment plants (referred to as wastewater surveillance) provides a representative view of infection in a particular area. Wastewater surveillance can be more economical and faster than clinical surveillance for understanding the prevalence of infectious diseases in a community and can lead to earlier detection (Kitajima et al., 2020; Murakami et al., 2020). Research has shown that concentration of severe acute respiratory syndrome coronavirus 2 (SARS-CoV-2) in wastewater foreshadows the number of clinically confirmed cases in the corresponding population, enabling earlier outbreak alerts (Ando et al., 2023; Kagami et al., 2023; Peccia et al., 2020; Tiwari et al., 2022). Although surveillance of wastewater discharged from facilities has been used to encourage testing and preventive behavior against infection for residents of those facilities (Betancourt et al., 2021; Kitajima et al., 2022), information based on surveillance at wastewater treatment plants has also been used to understand trends in infection status, particularly among experts (Dean, 2022). In Japan, since the reclassification (i.e., downgrading) of COVID-19’s legal status on May 8, 2023, infection have been tracked using weekly reports from selected sentinel medical institutions. This change has led to delays in understanding real-time infection status. Consequently, wastewater surveillance, which provides quicker insights (approximately 1–2 weeks earlier than sentinel surveillance), has gained prominence (Ando et al., 2023).

Current wastewater surveillance data are available on dedicated websites, such as those provided by the U.S. Centers for Disease Control and Prevention (Centers for Disease Control and Prevention, 2023) and the Netherlands’ Coronavirus Dashboard (Coronavirus Dashboard, 2023). In Japan, some local governments also share wastewater surveillance data (City of Sapporo, 2023b; Komatsu City, 2023). Few studies have examined public attitudes toward wastewater surveillance, with most respondents appreciating its utility (Holm et al., 2022; LaJoie et al., 2022; Takagi et al., 2023). However, the impact of this form of surveillance on the public interest in COVID-19 and its preventive behavior remain unclear.

Therefore, this study aimed to investigate how presenting wastewater surveillance data that reflected infection status influenced people’s perceptions and behavior related to COVID-19. We compared the reactions to information from sentinel surveillance alone with those from a combination of wastewater and sentinel clinical surveillance data. Specifically, we conducted a questionnaire survey to gauge participants’ understanding of, interest in, relief regarding, and intent to adopt preventive behavior after presenting the wastewater surveillance data. Additionally, we analyzed the participants’ online search behavior one-week post-survey, considering that people’s stated intentions may not always match their actions. We believe that their online search behavior could reflect the shifts promoted by wastewater data.

## 2. Methods

### 2.1. Ethics

This study was approved by the Ethics Committee of the Center for Infectious Disease Education and Research at Osaka University (approval number 2023CRER0612). All participants provided informed consent.

### 2.2. Surveys

This study was designed as a randomized controlled trial. Participants were randomly selected and divided into control (n = 10,000) and intervention (n = 10,000) groups out of approximately 50,000 active users of Yahoo’s crowdsourcing service (Yahoo! crowdsourcing) as of July 2023. These users were part of a larger pool of approximately one million registered users, all of whom were aged ≥15 years in Japan. An online survey was targeted for those aged ≥18 years and conducted for both groups between August 4 and August 7, 2023 until 1,000 valid responses were collected from each group.

The primary goal of this study was to understand the influence of providing information on wastewater surveillance, which reflected the infection status on individuals’ perceptions of and responses to COVID-19. The sample size was determined in consultation with Yahoo! JAPAN based on survey feasibility and a lack of prior knowledge regarding the potential impact of such information. With a population size of 125,000,000 in Japan, a sample size of approximately 400 is needed to achieve a 5% margin of error with a 95% confidence interval (Serdar et al., 2021). The sample size of this study surpassed that benchmark.

The questionnaires and information presented in the survey were written in Japanese. For both the control and intervention groups, data from more than 40% of the targeted participants were collected on August 4, and by August 5, the rate had exceeded 80% (Figure S1). Our survey coincided with the custom period in Japan when many people returned to their hometowns in mid-August. Many people took vacation days between August 11 (a national holiday) and 16. The participants assessed the infection status information using the URL provided (Figure 1). Those who answered “I did not read the content carefully” were excluded (27 in the control group and 36 in the intervention group). Participants who provided valid responses received a reward equivalent to 100 Japanese Yen.

**Figure 1.**
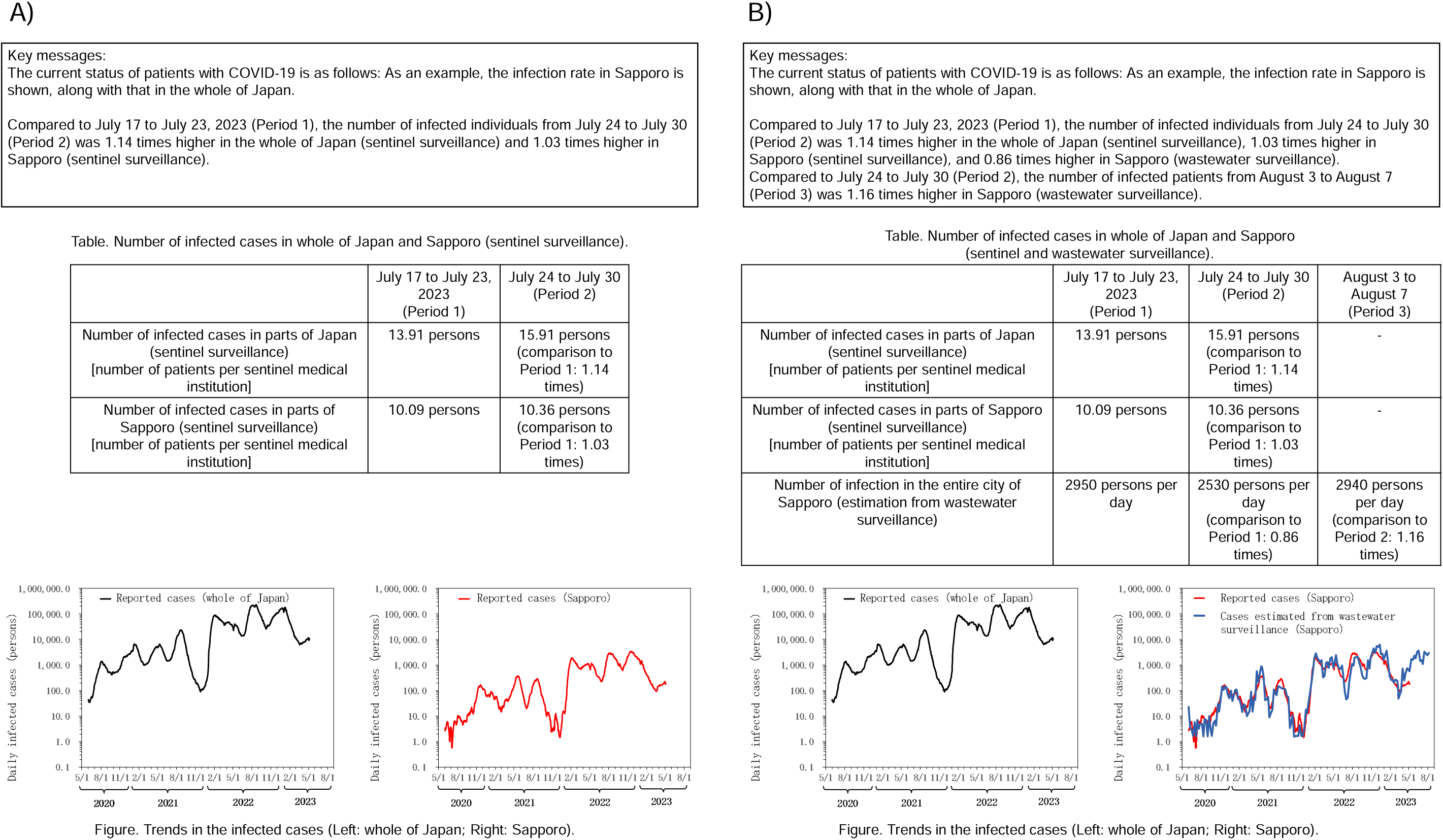
Information provided to participants. A) absence of wastewater surveillance information (control group); b) presence of wastewater surveillance information (intervention group).

The questionnaire comprised four major sections. Two authors [MM (specializing in risk science and environmental engineering) and SN (focusing on epidemiology and public health)] with expertise in risk perception and preventive behavior regarding COVID-19 (Adachi et al., 2022; Uchibori et al., 2022) were consulted to frame the questionnaire in the context of the COVID-19 pandemic. In the first section, prior to introducing any information on the infection status (covered in the second section), the participants rated their interest in COVID-19, changes in case numbers, their overall preventive behavior, mask usage, and attitudes toward vaccination using a five-point Likert scale (1 = strongly disagree to 5 = strongly agree). These evaluations were later labeled as “interest in COVID-19 (pre),” “interest in infection status (pre),” “overall preventive behavior (pre),” “mask-wearing (pre),” “awareness of the importance of COVID-19 vaccine (pre).”

Participants were then given different sets of information in the second section, depending on whether they were in the control or intervention group (Figure 1). The control group received the latest data on the numbers of reported cases from all of Japan and Sapporo (City of Sapporo, 2023a), including specific details from the two periods in July 2023 [July 17 to July 23, 2023 (Period 1), and July 24 to July 30 (Period 2)], along with a comparison of these periods. This information was shown both narratively and tabularly. Moreover, the number of daily reported cases from June 1, 2020 to May 7, 2023 in both Japan and Sapporo was represented by a figure.

The intervention group received information on wastewater surveillance in Sapporo, in addition to the number of cases described above. The wastewater surveillance estimated the number of cases in Sapporo for five days after sample collection based only on the SARS-CoV-2 concentration in wastewater (Ando et al., 2023). In addition to the data for Periods 1 and 2, the estimated figures from August 3 to August 7 (Period 3) and its ratio compared with Period 2 were also provided tabularly with a key message. The wastewater surveillance data estimated the total number of infected individuals in Sapporo from June 2020 to August 7, 2023, which was represented by a figure. This estimate was compared to the total confirmed cases from a notifiable clinical surveillance conducted throughout Japan and Sapporo until the downgrading of the legal status of COVID-19 (May 8, 2023), showing good agreement between the estimated and clinically confirmed cases in Sapporo.

Sapporo was chosen because of its capability to present estimates of the number of infected individuals (Ando et al., 2023) and because of their local government’s weekly updates on COVID-19 information based on wastewater surveillance on its website (City of Sapporo, 2023b).

In the third section, following the presentation of COVID-19 information in the second section and ensuring that the participant had read the content (which was confirmed with a “I read the content carefully” or “I did not read the content carefully”), respondents were prompted to indicate their understanding of, interest in, and relief regarding the COVID-19 infection status. They also provided their intentions for overall preventive behavior, mask-wearing, and vaccination as control measures using a five-point Likert scale (1 = strongly disagree to 5 = strongly agree).

These evaluations were subsequently labeled as “understanding of infection status (post),” “interest in infection status (post),” “relief regarding infection status (post),” “intent to engage in overall preventive behavior (post),” “intent to wear masks (post),” and “intent to receive COVID-19 vaccine (post).” In the fourth section, the participants responded to questions about their gender, age, place of residence (at the prefecture level), and history of COVID-19. There were no missing data for any of the survey items.

Next, we obtained population-based data on online search behavior from Yahoo! JAPAN. These data captured participants’ Yahoo! JAPAN search engine usage within one week before and after the online survey for both the control and intervention groups. Specifically, we first obtained information regarding the top 100 and bottom 100 keywords with the most significant search differences between the two groups post-survey. Based on this information, we established the following five lists of search keywords (terms in the keyword lists are indicated by double quotation marks):

(1) “bullet train” [*jr Hokkaido* or *jr East Japan* or *jr Tokai* or *jr West Japan* or *jr Shikoku* or *jr Kyushu* or *Shinkansen* (bullet train)]
(2) “highway” [*kousokudouro* (highway)]
(3) “airplane” [*ana* or *jal* or *Zennihonkuuyu* (All Nippon Airways) or *Zennikkuu* (an abbreviation of All Nippon Airways) or *Koukuu* (an abbreviation of Japan Airlines) or *hikouki* (airplane) or *kuukou* (airport)]
(4) “COVID-19” [*korona* (corona) or *covid*]
(5) “wastewater” [*gesui* (wastewater)].

Lists (1) and (3) represent bullet trains with their respective operators and airplane-related terms, including major Japanese airlines, respectively.

Considering the decreased travel in Japan during 2020 and 2021 because of the COVID-19 pandemic (Cabinet Office: Shogo Maeda, 2022), anticipating increased travel by mid-August 2023, after the downgrading of COVID-19’s legal status, we decided to use keyword lists (1) to (3). Keywords were extracted even when flanked by other characters [e.g., *koronauirusu* (i.e., coronavirus)] and were recognized irrespective of half-width or full-width characters, or case distinction.

Data regarding the search numbers within a week before and after the survey were obtained for both groups. If the search count was five or less, the exact number was withheld. Additionally, we acquired data on the ratio of the total Yahoo searchers in the week before and after the survey. The post-survey Yahoo user count was 1.0050 times greater than that in the pre-survey.

### 2.3. Statistical analysis

First, to ensure that the random assignment of the participants was done properly, we performed Z-pooled exact unconditional, chi-square, Fisher-Freeman-Halton, or Mann-Whitney’s U tests to investigate differences in “interest in COVID-19 (pre),” “interest in infection status (pre),” “overall preventive behavior (pre),” “mask-wearing (pre),” “awareness of the importance of COVID-19 vaccine (pre),” gender, age, prefecture of residence [Hokkaido (prefecture where Sapporo is located) or other prefectures], and history of COVID-19 infection between the control and intervention groups.

Next, we calculated Spearman’s correlation coefficients for the control and intervention groups among “understanding of infection status (post),” “interest in infection status (post),” “relief regarding infection status (post),” “intent to engage in overall preventive behavior (post),” “intent to wear masks (post),” and “intent to receive COVID-19 vaccine (post).” We then used Mann-Whitney’s U test to assess differences in “understanding of infection status (post),” “interest in infection status (post),” “relief regarding infection status (post),” “intent to engage in overall preventive behavior (post),” “intent to wear masks (post),” and “intent to receive COVID-19 vaccine (post)” between the control and intervention groups. Furthermore, to examine how participants’ psychological factors had additional effect of the intervention on the outcomes, we performed ordinal logistic regression analyses for “interest in infection status (post),” “relief regarding infection status (post),” and “intent to engage in overall preventive behavior (post)” that showed the significant differences in the Mann-Whitney’s U test above. The intervention, “interest in COVID-19 (pre),” “overall preventive behavior (pre),” and their interaction terms with the intervention were used as explanatory variables. “Interest in COVID-19 (pre)” and “overall preventive behavior (pre)” were centered. The variance inflation factor was 1.00 for the intervention and 3.06–3.43 for the other variables, indicating that multicollinearity was sufficiently small.

Finally, we compared the proportion of searchers for each keyword list between the control and intervention groups in the week before and after responding to the questionnaire survey. For this purpose, we used a Z-pooled exact unconditional test. This test was chosen because its appropriateness in evaluating associations in two × two tables, especially when dealing with a low number of searchers (Mehrotra et al., 2003). Analyses were conducted using IBM SPSS 28 (IBM, Chicago, IL, USA) or R (Calhoun, 2022; R Development Core Team, 2021).

## 3. Results

Table 1 shows the differences in the characteristics of participants in the control and intervention groups before the presentation of information on infection status. There were no significant differences (*P* > 0.05) in individual attributes such as gender, age, prefecture of residence, history of COVID-19, or in interest in and preventive behavior regarding COVID-19 before the presentation of information.

**Table 1.** Participants’ characteristics. N: number of participants; SD: standard deviation.

| | | N (%) or *mean (SD)* | | $\phi$, Cramer's V, or *r* | *P* |
| --- | --- | --- | --- | --- | --- |
|  |  | Control | Intervention |  |  |
| Gender | Woman | 276 (27.6%) | 269 (26.9%) | 0.033 | 0.571 |
|  | Man | 717 (71.7%) | 719 (71.9%) |  |  |
|  | Other | 1 (0.1%) | 4 (0.4%) |  |  |
|  | Don't answer | 6 (0.6%) | 8 (0.8%) |  |  |
| Age | 18–29 | 38 (3.8%) | 30 (3.0%) | 0.061 | 0.396 |
|  | 30–39 | 116 (11.6%) | 100 (10.0%) |  |  |
|  | 40–49 | 265 (26.5%) | 294 (29.4%) |  |  |
|  | 50–59 | 331 (33.1%) | 348 (34.8%) |  |  |
|  | 60–69 | 167 (16.7%) | 166 (16.6%) |  |  |
|  | 70–79 | 73 (7.3%) | 53 (5.3%) |  |  |
|  | 80 and more | 4 (0.4%) | 4 (0.4%) |  |  |
|  | Don't answer | 6 (0.6%) | 5 (0.5%) |  |  |
| Prefecture | Hokkaido | 39 (3.9%) | 43 (4.3%) | −0.010 | 0.671 |
|  | Other | 961 (96.1%) | 957 (95.7%) |  |  |
| Covid history | Yes | 193 (19.3%) | 189 (18.9%) | 0.010 | 0.901 |
|  | No | 803 (80.3%) | 808 (80.8%) |  |  |
|  | Don't answer | 4 (0.4%) | 3 (0.3%) |  |  |
| Interest in and preventive behavior against COVID-19 before providing information | Interest in COVID-19 (pre) | 3.51 (1.04) | 3.52 (1.06) | 0.010 | 0.666 |
|  | Interest in infection status (pre) | 3.25 (1.15) | 3.32 (1.15) | 0.028 | 0.209 |
|  | Overall preventive behavior (pre) | 3.80 (1.00) | 3.82 (0.95) | 0.001 | 0.968 |
|  | Mask-wearing (pre) | 3.69 (1.32) | 3.66 (1.34) | −0.011 | 0.616 |
|  | Awareness of the importance of COVID-19 vaccine (pre) | 3.40 (1.24) | 3.41 (1.19) | −0.004 | 0.857 |

There was no significant association between relief regarding infection status and interest in or intention for preventive behavior in either the control or intervention groups after the information presentation, except for the association between relief regarding infection status (post) and intent to receive COVID-19 vaccine (post) in the intervention group (Table S1; *P* > 0.05). By contrast, significant associations were observed between interest in and intention for preventive behavior (*P* < 0.05). For example, the Spearman’s correlation coefficient between interest in infection status (post) and intent to engage in overall preventive behavior (post) in the intervention group was 0.639 (*P* < 0.001).

Table 2 shows differences in understanding of, interest in, relief regarding, and intention for preventive behavior regarding COVID-19 between the two groups after information presentation. Interest in infection status (post), relief regarding infection status (post), and intent to engage in overall preventive behavior (post) in the intervention group were significantly higher than those in the control group (*P* < 0.05; *r* = 0.045–0.048). No significant differences were found in understanding of infection status (post), intent to wear masks (post), and intent to receive COVID-19 vaccine (post) (*P* > 0.05).

**Table 2.**
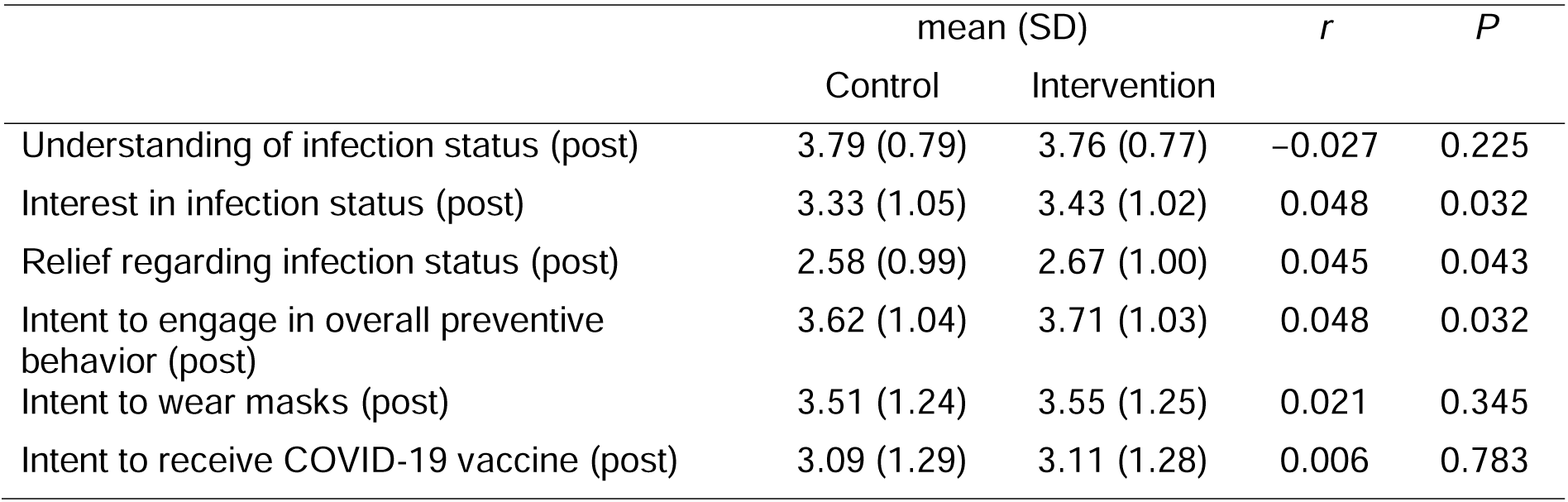
Differences in understanding of, interest in, relief regarding, and intention for preventive.

|  | mean (SD) |  | *r* | *P* |
| --- | --- | --- | --- | --- |
|  | Control | Intervention |  |  |
| Understanding of infection status (post) | 3.79 (0.79) | 3.76 (0.77) | −0.027 | 0.225 |
| Interest in infection status (post) | 3.33 (1.05) | 3.43 (1.02) | 0.048 | 0.032 |
| Relief regarding infection status (post) | 2.58 (0.99) | 2.67 (1.00) | 0.045 | 0.043 |
| Intent to engage in overall preventive behavior (post) | 3.62 (1.04) | 3.71 (1.03) | 0.048 | 0.032 |
| Intent to wear masks (post) | 3.51 (1.24) | 3.55 (1.25) | 0.021 | 0.345 |
| Intent to receive COVID-19 vaccine (post) | 3.09 (1.29) | 3.11 (1.28) | 0.006 | 0.783 |

Table 3 shows the associations of interest in COVID-19 (pre), overall preventive behavior (pre), intervention, and their interaction terms with interest in infection status (post), relief regarding infection status (post), and intent to engage in overall preventive behavior (post). All outcomes were significantly and positively associated with the intervention (*P* < 0.05). Interest in COVID-19 (pre) and overall preventive behavior (pre) were significantly and positively associated with interest in infection status (post) and intent to engage in overall preventive behavior (post) (*P* < 0.05), whereas their interaction terms with intervention were not significantly associated with these two outcomes (*P* > 0.05). In contrast, overall preventive behavior (pre) and the interaction term with intervention were significantly negatively and positively associated with relief regarding infection status (post), respectively (*P* < 0.05). Interest in COVID-19 (pre) was not significantly associated with relief regarding infection status (post) (*P* > 0.05), but its interaction term with intervention showed a significant negative association (*P* < 0.05).

**Table 3.** Associations of intervention, interest (pre), overall preventive behavior (pre), and their interaction terms with interest (post), relief (post), and intent to engage in overall preventive behavior against COVID-19 (post). VIF: variance inflation factor; aOR: adjusted odds ratio; CI: confidence interval.

|  | VIF | Interest in infection status (post) | aOR (95% CI)<br>Relief regarding infection status (post) | Intent to engage in overall preventive behavior (post) |
| --- | --- | --- | --- | --- |
| Intervention | 1.00 | 1.23 (1.05–1.45) | 1.17 (1.00–1.38) | 1.29 (1.08–1.53) |
| Interest in COVID-19 (pre) | 3.40 | 2.91 (2.50–3.38) | 1.06 (0.92–1.22) | 2.03 (1.74–2.36) |
| Overall preventive behavior (pre) | 3.16 | 1.85 (1.59–2.15) | 0.85 (0.73–0.98) | 4.02 (3.41–4.74) |
| Intervention * Interest in COVID-19 (pre) | 3.29 | 0.94 (0.77–1.15) | 0.80 (0.66–0.97) | 0.83 (0.68–1.02) |
| Intervention * Overall preventive behavior (pre) | 3.06 | 0.87 (0.70–1.08) | 1.24 (1.01–1.53) | 1.17 (0.94–1.46) |

Table 4 shows the proportion of participants who engaged in search behavior after the survey was administered. The proportion of participants who searched for “bullet train” was marginally higher in the intervention group than in the control group, although the difference was not significant (*P* = 0.129; φ = 0034). When substituting the number three for “highway” (i.e., 1/2 of the upper limit rounded to the nearest integer) in the control group, the intervention group had a significantly higher proportion of searchers (*P* < 0.05; φ = 0048). However, using the upper limit of five, the difference was not significant (*P* = 0.146; φ = 0034). The proportions of participants who searched for “airplane” and “COVID-19” were similar between the two groups (*P* > 0.05). The number of participants who searched for “wastewater” was ≤5 for each group.

**Table 4.** Differences in proportions of keyword searches after providing information. N: number of participants.

| | N (%) | | $\varphi$ | $P$ |
| --- | --- | --- | --- | --- |
|  | Control | Intervention |  |  |
| "Bullet train" | 21 (2.1%) | 32 (3.2%) | 0.034 | 0.129 |
| "Highway" | ≤5 (≤0.5%) | 11 (1.1%) | 0.034<sup>a</sup><br>(0.048)<sup>b</sup> | 0.146<sup>a</sup><br>(0.034)<sup>b</sup> |
| "Airplane" | 47 (4.7%) | 37 (3.7%) | -0.025 | 0.273 |
| "COVID-19" | 16 (1.6%) | 17 (1.7%) | 0.004 | 0.915 |
| "Wastewater" | ≤5 (0.5%) | ≤5 (0.5%) | - | - |

<sup>a</sup> "5" (i.e., the upper limit) was used for the number of control group.
<sup>b</sup> "3" (i.e., 1/2 of the upper limit rounded to the nearest integer) was used for the number of control group.

Table S2 shows the proportion of participants who searched for keyword lists during the week before the survey. There was no significant difference in the proportion of participants who searched for any of the keyword lists between the two groups (*P* > 0.05).

## 4. Discussion

Using a randomized controlled trial design, this study investigated whether the presentation of information on COVID-19 infection status based on wastewater surveillance increased interest in, relief regarding, and intention for preventive behavior against COVID-19. We also analyzed changes in online search behavior after the provision of the aforementioned information.

The participants in this study did not differ in gender, age, place of residence, history of COVID-19 infection, and interest in and preventive behavior against COVID-19 prior to information presentation. Similarly, the proportion of participants who searched for each keyword list prior to the survey was consistent between the control and intervention groups. This confirmed that the randomization assignment of the participants was executed well based on the observed variables.

Next, the presentation of information on infection status based on wastewater surveillance increased interest in, relief regarding, and intention for overall preventive behavior against COVID-19. This finding underscores the utility of such information. Furthermore, this information presentation may have caused a slight uptick in search behavior related to highways as a mode of transportation. However, the difference between the control and intervention groups was not clearly specified, because the number of participants who searched was below the upper limit in the control group. In addition, although not significant, there was a trend toward more searches related to bullet trains in the intervention group. Such spikes in search behavior may imply an increase in return travel during the mid-August vacation season.

Traffic volume on highways from August 9 to August 16, 2023 (approximately one week post-survey) was 1.07 times that in 2022 (NEXCO EASTTOP, 2023). On August 10, 2023, potential nationwide cancellations of bullet trains owing to Typhoon No. 7, including JR Tokai bullet trains connecting major cities, were announced. Subsequently, there were cancellations and delays between August 15 and 17, 2023. The passenger count on the JR Tokai bullet train from August 10 to August 17 was 1.25 times that in 2022 (JR Central Japan Railway Company (JR Toukai), 2023). Therefore, after the survey, it is likely that participants who planned to return or travel during the mid-August vacation season began searching for highway and bullet train-related terms. The wastewater surveillance data presentation might have nudged people toward pre-COVID-19 routine behavior, such as using highways or bullet trains for travel, along with a rise in interest in, relief regarding, or intention for preventive behavior against COVID-19.

It is challenging to interpret why wastewater surveillance-based information intensified interest in, relief regarding, and intention toward overall COVID-19 preventive behavior, and why there was a spike in search behavior related to highways. One of the assumptions underlying the utility of presenting wastewater surveillance-based information is that people generally have a favorable perception of the wastewater surveillance approach (Holm et al., 2022; LaJoie et al., 2022; Takagi et al., 2023).

This study also indicated that significant associations existed between interest in and intention for overall preventive behavior post-presentation. However, no significant associations were observed between relief regarding and either interest in or intention for overall preventive behavior. Results from the ordinal logistic regression analysis showed varied associations with explanatory variables between relief and the interest or intention post-presentation. The association between interest and intention aligns well with the finding in a previous study (Yamagata et al., 2023). The present study suggests that participants who experienced increased interest and intention toward overall preventive measures for COVID-19 owing to the presentation of wastewater surveillance information differed in characteristics from those who felt an enhanced sense of relief regarding the infection status.

Interest in infection status (post) or intent to engage in overall preventive behavior (post) was significantly and positively associated with interest in COVID-19 (pre) and overall preventive behavior (pre) but not with the interaction items with intervention. This means that although participants who had previously shown more interest in COVID-19 and engaged in preventive behavior were more interested in the infection status or intended to engage in overall preventive behavior post-survey, the presentation of wastewater surveillance-based information had no additional effect on these outcomes.

In contrast, relief regarding infection status (post) was negatively associated with overall preventive behavior (pre) and the interaction term between intervention and interest in COVID-19 (pre). It was also positively associated with the interaction term between the intervention and overall preventive behavior (pre). This suggests that those who had engaged in preventive behavior against COVID-19 were more likely to be concerned about its infection status even after receiving information on the infection status, but the wastewater surveillance data increased their sense of relief. In addition, individuals who were less interested in COVID-19 felt more relief when they received wastewater surveillance information.

The data from wastewater surveillance highlighted in this study provided two primary advantages: first, it allows for early detection as wastewater surveillance information captures more recent infection status compared to clinical surveillance data; second, it ensures representativeness by enabling comparisons with infection status derived from a comprehensive survey conducted until the downgrading of the legal status of COVID-19 (Kitajima et al., 2020; Murakami et al., 2020).

The infection status highlighted by the wastewater surveillance data in this study was nearly at its peak; however, it showed no significant fluctuations in the count of infections over the previous 2–3 weeks. This stability may have offered some individuals, particularly those who were less interested in COVID-19, a heightened sense of relief. Their relief may have stemmed from the perceived absence of worsening conditions, validating their indifference. Alternatively, knowledge of this novel surveillance method may have provided comfort. Using wastewater for infection tracking is a new approach, but its public awareness remains limited (Holm et al., 2022). Those practicing COVID-19 preventive measures may find reassurance from these data, viewing it as an additional resource for informing their actions.

This study has some limitations. First, although we confirmed that there were no significant differences in individual attributes, such as gender and age, and psychological factors between the control and intervention groups prior to presenting the information, we were unable to verify differences in economic status and occupation. Second, because the participants were exclusively registered active Yahoo crowdsourcing users, caution is necessary when generalizing the findings. Third, the impact of presenting wastewater surveillance information may vary depending on the infection status. Our study focused on a scenario in which the infection numbers were at their peak, but there were not significant fluctuations in the previous 2–3 weeks. Hence, longitudinal monitoring of the perceptions and behavior of individuals exposed to this information is essential to understand the impact of different infection statuses. Fourth, regarding the online search of the keyword list, we could not determine the actual intent of the participants. Furthermore, some extracted searches might not have aligned with the study’s intended focus because keywords were included even if they contained characters before or after the targeted keywords. We assumed that the increase in the number of participants who conducted online searches was associated with keywords such as “highways” or “bullet trains.” Future studies should combine online search analyses with behavioral observations. Fifth, the effects of this study, which focused on infection status information derived from wastewater surveillance, showed only a minimal impact on preventive behavior and increased search behavior related to highways. Further research is required to develop more impactful information presentation methods.

## 5. Conclusions

Using a randomized controlled trial design, this study investigated the perceptions and intentions regarding preventive behavior related to COVID-19 by presenting infection status information from wastewater surveillance. Furthermore, we analyzed the behavioral changes brought about by this information using online search patterns. This is the first study to reveal the effects of wastewater surveillance information on the perceptions of and responses to COVID-19. The main findings of this study are as follows:

- Presentation information on infection status via wastewater surveillance increased the interest in, relief regarding, and intention for overall preventive behavior regarding COVID-19. However, there was no increase in the understanding of the infection status and intention to wear masks and receive the COVID-19 vaccination.
- The characteristics of the participants, where the presentation of wastewater surveillance information triggered a further rise in interest or intention for overall preventive behavior

regarding COVID-19, varied from those feeling an enhanced sense of relief regarding the infection status. The additional effect of presenting information on interest or intention for overall preventive behavior was unaffected by prior interest in COVID-19 or overall preventive behavior. In contrast, a heightened sense of relief was more prominent among those who already practiced COVID-19 preventive behavior or those who were less interested in COVID-19.

- The presentation of wastewater surveillance information may have led to a slight uptick in online searches, such as for highways pertinent to homecoming during the mid-August vacation season, suggesting a possible resumption of habits from the pre-COVID-19 times.
- The utility of presenting wastewater surveillance information was demonstrated through observations of changes in people’s perceptions of and response to infections.

## Declaration of Competing Interest

Michio Murakami reports a relationship with NJS CO., LTD. that includes: consulting or advisory. Masaaki Kitajima reports a relationship with Shionogi & Co., Ltd. that includes: funding grants. Masaaki Kitajima reports a relationship with AdvanSentinel, Inc. that includes: funding grants. Masaaki Kitajima has patent pending to Shionogi & Co., Ltd. Other authors declared no competing interest.

## Author contributions

MM: Conceptualization, Methodology, Formal analysis, Investigation, Visualization, Funding acquisition, Writing – original draft.

SN: Conceptualization, Methodology, Funding acquisition, Writing – review & editing. HA: Resources, Writing – review & editing.

MK: Resources, Funding acquisition, Writing – review & editing.

## Data Availability

We have included all the results produced in the present work in the manuscript. Note that the raw data used in the study were provided by Yahoo! JAPAN, as described in this paper. We are unable to attach all the raw data for each participant in this paper due to the contract and ethical restrictions.

## Acknowledgements

We would like to thank Editage (www.editage.com) for English language editing and Yahoo! JAPAN for the survey. We are also grateful for helps: Ms. Aya Nishida (Osaka University).

## Funding

This work was supported by “The Nippon Foundation - Osaka University Project for Infectious Disease Prevention,” a grant PRESTO [JPMJPR22R8] from the Japan Science and Technology Agency, and the Japan Agency for Medical Research and Development (AMED) under grant number JP22fk0108508. The funders had no role in study design, data collection and analysis, decision to publish, or preparation of the manuscript.

## Statements

During the preparation of this manuscript, the authors used DeepL solely for the purpose of the possible improvement of English language expression. The authors created the original texts before using this tool. The authors reviewed and edited the content as needed, after using this tool. Furthermore, the paper was carefully edited by native professional editors. The authors take full responsibility for the content of the publication.

## Notes

### Author Declarations

The Ethics Committee of the Center for Infectious Disease Education and Research at Osaka University gave ethical approval for this work (approval number 2023CRER0612).

