## Supplementary File for "Perceptions and responses to COVID-19 through wastewater surveillance information and online search behavior: A randomized controlled trial"

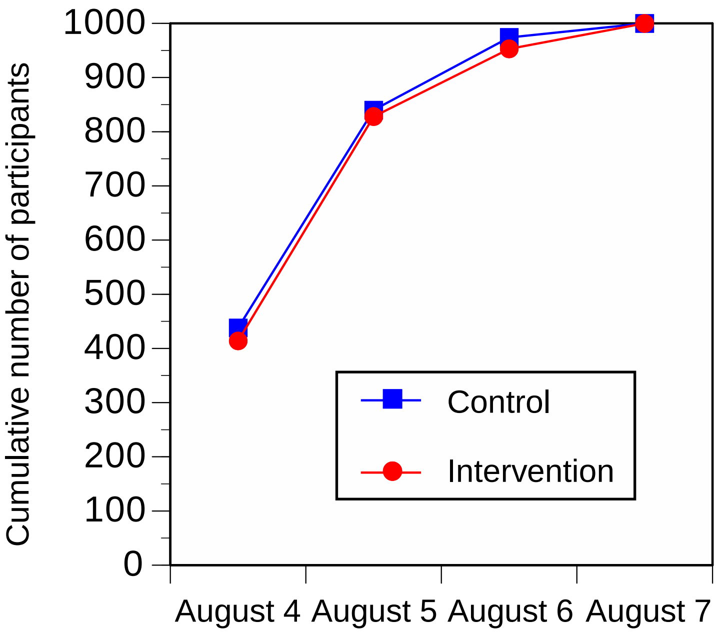


Figure S1. Cumulative number of participants.

Table S1. Spearman’s correlation among understanding of, interest in, relief regarding, and intention for preventive behavior against COVID-19 after providing information.

| | | Upper right: control | | | | | |
|---|---|---|---|---|---|---|---|
| | | Understanding of infection status (post) | Interest in infection status (post) | Relief regarding infection status (post) | Intent to engage in overall preventive behavior (post) | Intent to wear masks (post) | Intent to receive COVID-19 vaccine (post) |
| Lower left: intervention | Understanding of infection status (post) | - | 0.461** | 0.139** | 0.339** | 0.265** | 0.269** |
| | Interest in infection status (post) | 0.523** | - | 0.009 | 0.692** | 0.592** | 0.509** |
| | Relief regarding infection status (post) | 0.111** | 0.044 | - | −0.056 | −0.060 | 0.030 |
| | Intent to engage in overall preventive behavior (post) | 0.404** | 0.639** | −0.027 | - | 0.786** | 0.574** |
| | Intent to wear masks (post) | 0.275** | 0.581** | −0.057 | 0.784** | - | 0.524** |
| | Intent to receive COVID-19 vaccine (post) | 0.295** | 0.500** | 0.076* | 0.502** | 0.491** | - |

** $P < 0.01$, * $P < 0.05$

Table S2. Differences in proportions of keyword searches before providing information. N: number of participants.

| | N (%) | | $\varphi$ | *P* |
|---|---|---|---|---|
| | Control | Intervention | | |
| "Bullet train" | 15 (1.5%) | 15 (1.5%) | 0.000 | 1.000 |
| "Highway" | ≤5 (≤0.5%) | ≤5 (≤0.5%) | - | |
| "Airplane" | 37 (3.7%) | 29 (2.9%) | −0.022 | 0.345 |
| "COVID-19" | 18 (1.8%) | 18 (1.8%) | 0.000 | 1.000 |
| "Wastewater" | ≤5 (0.5%) | ≤5 (0.5%) | - | - |
